# Low coverage whole genome sequencing of low-grade dysplasia strongly predicts colorectal cancer risk in ulcerative colitis

**DOI:** 10.1101/2024.07.08.24309811

**Authors:** Ibrahim Al Bakir, Kit Curtius, George D Cresswell, Heather E Grant, Nadia Nasreddin, Kane Smith, Salpie Nowinski, Qingli Guo, Hayley L Belnoue-Davis, Jennifer Fisher, Theo Clarke, Christopher Kimberley, Maximilian Mossner, Philip D Dunne, Maurice B Loughrey, Ally Speight, James E East, Nicholas A Wright, Manuel Rodriguez-Justo, Marnix Jansen, Morgan Moorghen, Ann-Marie Baker, Simon J Leedham, Ailsa L Hart, Trevor A Graham

## Abstract

Patients with inflammatory bowel disease (IBD) are at increased risk of colorectal cancer (CRC), and this risk increases dramatically in those who develop low-grade dysplasia (LGD). However, there is currently no accurate way to risk-stratify patients with LGD, leading to both over- and under-treatment of cancer risk. Here we show that the burden of somatic copy number alterations (CNAs) within resected LGD lesions strongly predicts future cancer development. We performed a retrospective multi-centre validated case-control study of n=122 patients (40 progressors, 82 non-progressors, 270 LGD regions). Low coverage whole genome sequencing revealed CNA burden was significantly higher in progressors than non-progressors (p=2x10^-6^ in discovery cohort) and was a very significant predictor of CRC risk in univariate analysis (odds ratio = 36; p=9x10^-7^), outperforming existing clinical risk factors such as lesion size, shape and focality. Optimal risk prediction was achieved with a multivariate model combining CNA burden with the known clinical risk factor of incomplete LGD resection. The measurement of CNAs in LGD lesions is a robust, low-cost and rapidly translatable predictor of CRC risk in IBD that can be used to direct management and so prevent CRC in high-risk individuals whilst sparing those at low-risk from unnecessary intervention.

## Introduction

The management of colitis associated colorectal cancer (CA-CRC) risk suffers from both chronic under- and over-diagnosis and treatment. Patients with colitis are offered enrolment into endoscopic surveillance programs to detect early signs of cancer, namely the pathological diagnosis of dysplasia. Patients with low-grade dysplasia (LGD), itself a contentious definition^1–3^, have an approximately 30% chance of progressing to CRC at 10 years^4^ and this level of risk is considered sufficient to offer prophylactic colon resection. Major surgery has many risks, and subsequent detrimental impact on quality-of-life due to an ileal-pouch anal anastomosis or a stoma. Alternatively, patients can be offered watchful-waiting, where frequent colonoscopy and worry about CRC risk also severely impact quality-of-life. Patients and clinicians can have disparate thresholds for tolerable cancer risk^5,6^, and so the joint decision-making process of which route to follow can prove challenging. Ultimately, these decisions are made against the backdrop where more than two thirds of colitis patients with resected LGD are not actually at elevated risk of developing CRC, and so any treatment for these patients is over-treatment. Consequently, there is a major unmet need for accurate cancer risk stratification in colitis patients with LGD.

Some level of cancer risk stratification of colitis-associated LGD lesions is achieved from assessing clinical and endoscopic factors. Lesion size greater than 10mm, non-polypoid lesion shape or endoscopically invisible, and a previous history of indefinite dysplasia, are significantly associated with LGD progression to advanced neoplasia (namely high-grade dysplasia and CRC)^7^. We recently found that four clinicopathological variables, endoscopically visible LGD >1 cm, incomplete or impossible endoscopic resection, moderate/severe histological inflammation within 5 years of LGD diagnosis, and multifocal dysplasia provided excellent negative predictive value of cancer risk^8^. Other clinical risk factors known to be associated with progression to CRC in the general IBD population include a concomitant diagnosis of primary sclerosing cholangitis (PSC), cumulative inflammation burden, stricturing, the presence of inflammatory pseudopolyps and colonic scarring^9^.

The inflammatory micro-environment generated by colitis drives carcinogenesis through genomic and molecular pathway alterations that differ from those seen in sporadic CRC^10–13^. One key difference is the earlier onset of aneuploidy in colitis, which can be detected even in non-neoplastic colitic epithelium^13,14^. Studies that aim to exploit these molecular differences for risk stratification of LGD remain limited by the small sample sizes^15–17^, their use of “high risk” flat or invisible dysplasia lesions, and their reliance on flow cytometry and comparative genomic hybridisation, with further limitations in terms of the large quantities of preserved cells or DNA required.

Low pass whole genome sequencing (lpWGS) is a cost-efficient approach that utilises next generation sequencing to reliably detect copy number alterations (CNAs). The input can be minimal amounts of DNA (as low as 500 picograms), it is compatible with the highly degraded DNA that is typically derived from formalin-fixed paraffin-embedded (FFPE) blocks in hospital archives^18^, and it does not require a matched patient-specific reference genome for copy number estimation. In this study, we show that CNAs detected in lpWGS data from LGD biopsies of IBD patients very strongly predict future risk of high-grade dysplasia (HGD) and/or CRC, and that this genomic measurement adds significant prognostic value to the currently used clinicopathological variables.

## Results

### Copy number profiles can be derived from archival LGD samples by low pass whole genome sequencing

We conducted a retrospective case-control study (Figure 1) in which “progressors” (cases) were defined as IBD patients where LGD was the highest histological grade detected at colonoscopy, and who subsequently developed HGD or CRC within 5 years. The “non-progressors” (controls) were defined as IBD patients where LGD was the highest histological grade detected at colonoscopy, and who subsequently remained free from both HGD and CRC for at least 5 years.

**Figure 1:**
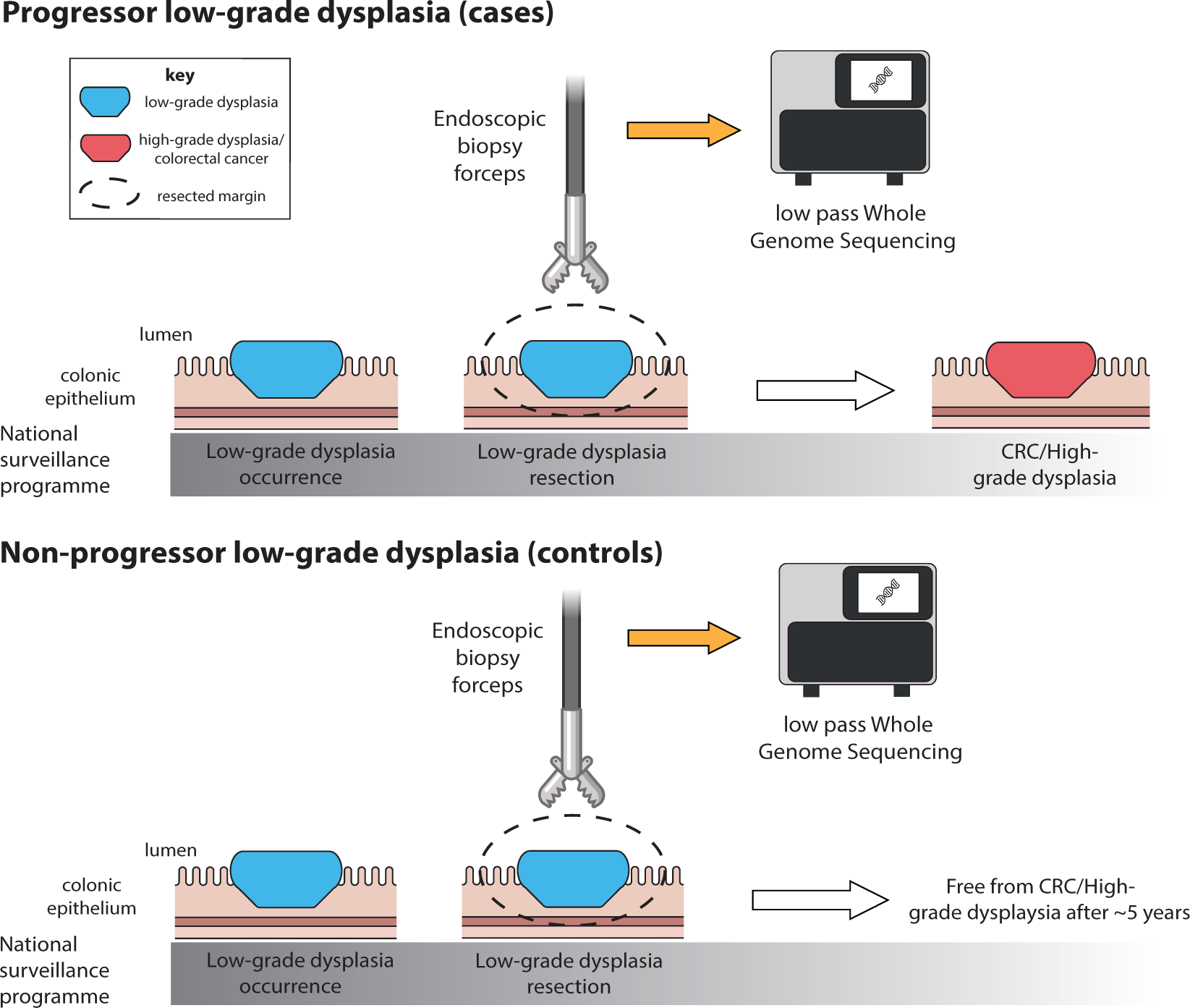
Study design. Schematic showing the detection and removal of low-grade dysplasia during routine IBD surveillance, with progressors developing high-grade dysplasia or colorectal cancer within 5 years (top), and non-progressors remaining free from these (bottom).

We identified a discovery cohort of 67 patients (22 “progressors” who developed HGD/CRC a median 427 days after LGD diagnosis, and 45 “non-progressors”) bearing 78 LGD lesions (see Table 1 and Methods for details). While progressors and non-progressor patients in the discovery cohort were well-matched by age, gender, duration of disease, PSC status and lesion location and morphology, the progressor cohort is statistically over-represented by larger lesions. A validation cohort of 55 patients was collected from University of Newcastle (n=15), University of Belfast (n=4), and Oxford University (n=36) that included a total of 59 LGD lesions (see Table 2).

**Table 1.** Summary of patients and LGD samples used as the discovery data by progression status, clinical features and endoscopic findings. *Fisher’s exact test, **Mann-Whitney test, *** X^2^ test of independence, NS = Not significant

| Data type | Variable | Progressor LGD | Non-progressor LGD | Significance between groups |
| --- | --- | --- | --- | --- |
| **Cases** | Number of patients | 22 patients | 45 patients | N/A |
|  | Number of lesions | 30 lesions | 48 lesions |  |
|  | Number of regions | 64 regions | 100 regions |  |
| **Clinical features** | Days to censor | 427 days<br>(i.e. developed HGD/CRC 1.2 years later)<br>*IQR 218 – 907 days* | 2,772 days<br>(i.e. CRC/HGD-free 7.6 years later)<br>*IQR 2,201 – 3,212 days* | N/A |
|  | Gender (% male) | 59% | 73% | NS*<br>(p = 0.27) |
|  | Median age (years) | 58<br>(range 34-77 years) | 60<br>(range 28-78 years) | NS**<br>(p = 0.30) |
|  | Median duration of disease (years) | 18<br>(range 8-48 years) | 27<br>(range 1-57 years) | NS**<br>(p = 0.19) |
|  | Proportion with PSC | 18.2% | 4.4% | NS*<br>(p = 0.09) |
| **Lesion features** | Lesion spatial distribution | 63% left colorectum<br>37% right colon | 69% left colorectum<br>31% right colon | NS*<br>(p = 0.63) |
|  | Lesion morphology | 38% polypoid<br>38% non-polypoid<br>24% invisible<br>n=29 | 54% polypoid<br>31% non-polypoid<br>15% invisible | NS***<br>(p = 0.34) |
|  | Median size of visible lesions (mm) | 20<br>(range 7-55)<br>n=23 | 8<br>(range 3-65)<br>n=41 | Significant**<br>(p = 0.0004) |

**Table 2.**
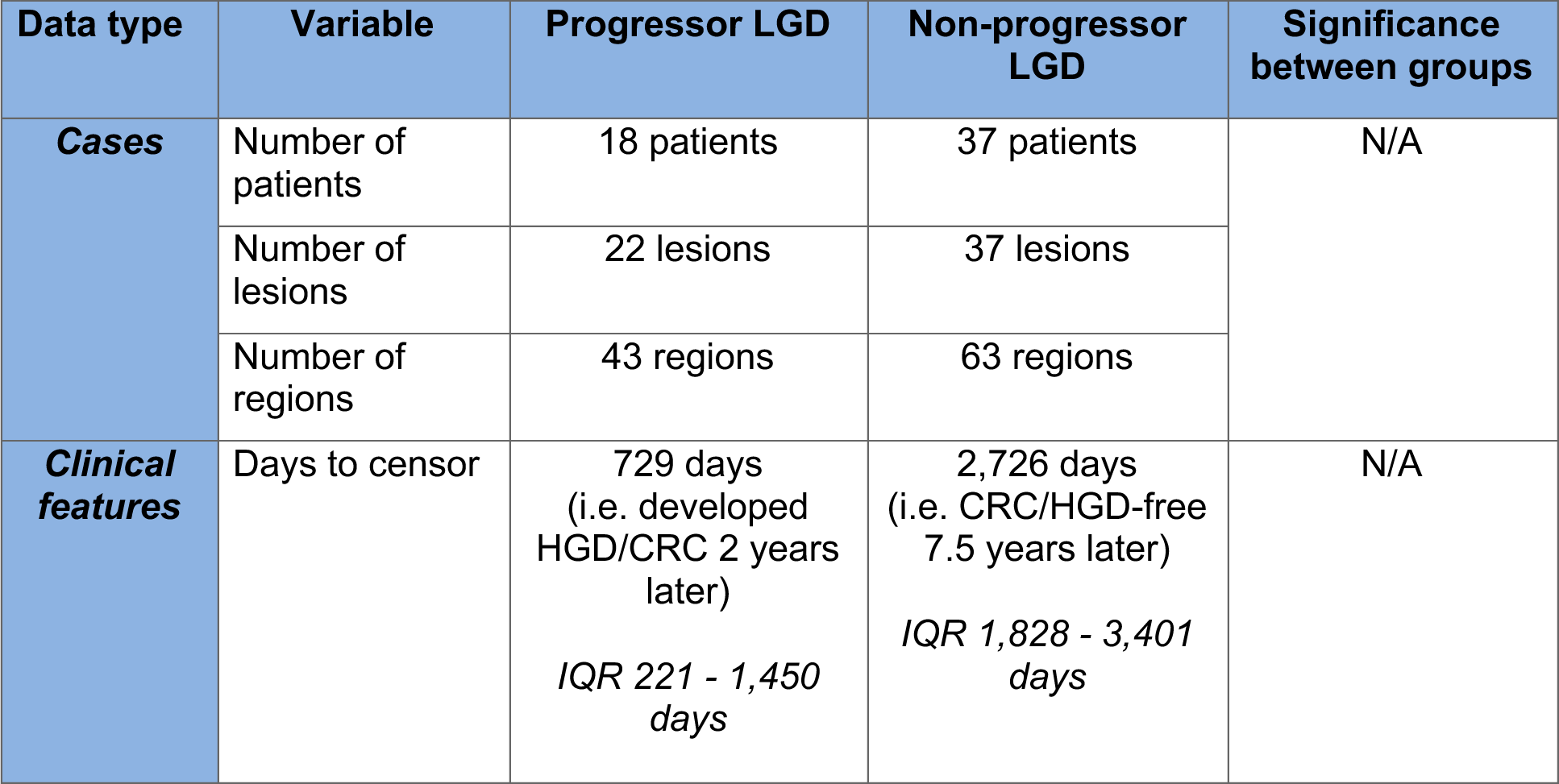
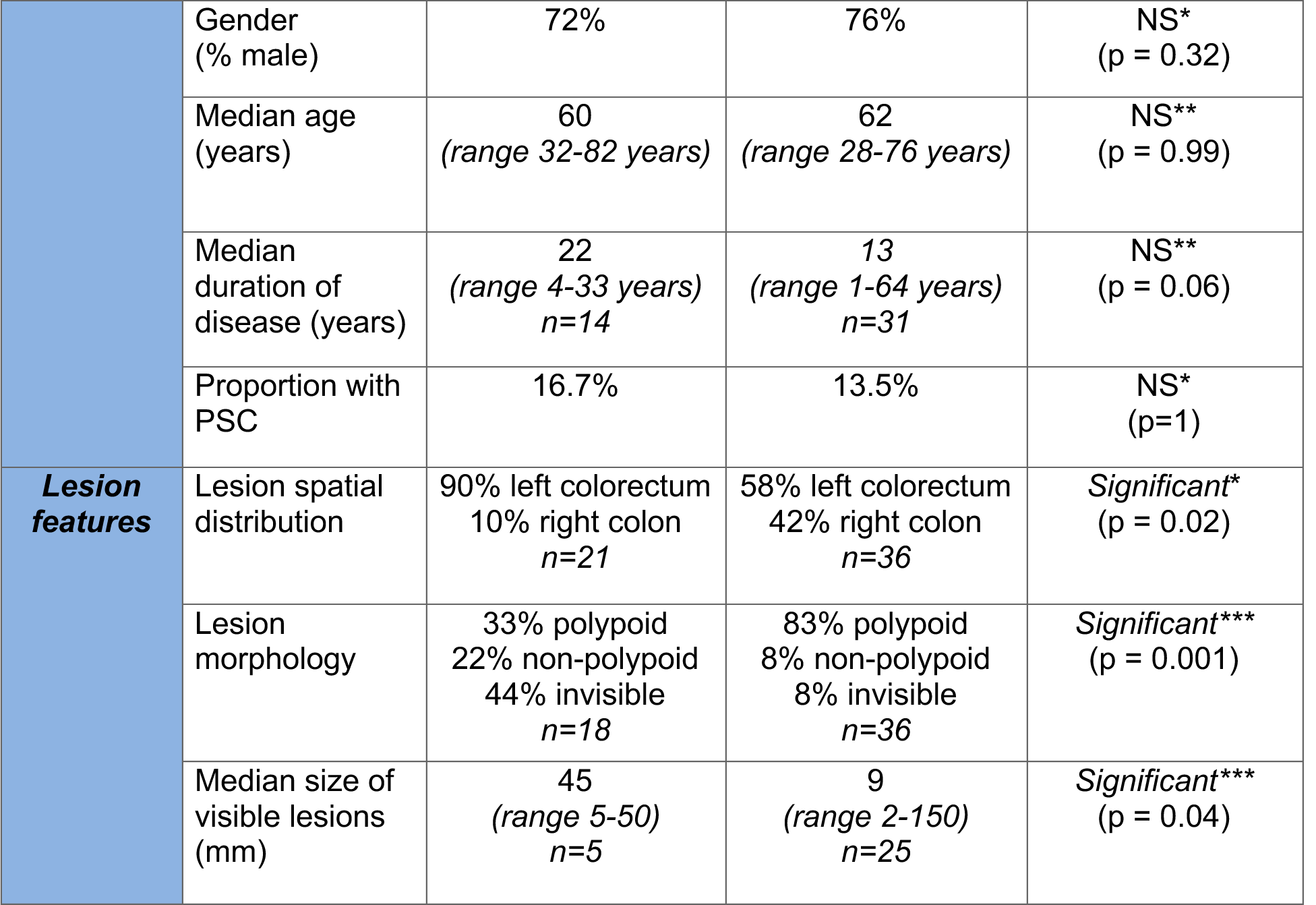
Summary of patients and LGD samples used as the validation data by progression status, clinical features and endoscopic findings. *Fisher’s exact test, **Mann-Whitney test, ***X^2^ test of independence, NS = Not significant

| Data type | Variable | Progressor LGD | Non-progressor LGD | Significance between groups |
| --- | --- | --- | --- | --- |
| **Cases** | Number of patients | 18 patients | 37 patients | N/A |
|  | Number of lesions | 22 lesions | 37 lesions |  |
|  | Number of regions | 43 regions | 63 regions |  |
| **Clinical features** | Days to censor | 729 days<br>(i.e. developed HGD/CRC 2 years later)<br>IQR 221 - 1,450 days | 2,726 days<br>(i.e. CRC/HGD-free 7.5 years later)<br>IQR 1,828 - 3,401 days | N/A |
|  | Gender (% male) | 72% | 76% | NS*<br>(p = 0.32) |
|  | Median age (years) | 60<br>(range 32-82 years) | 62<br>(range 28-76 years) | NS**<br>(p = 0.99) |
|  | Median duration of disease (years) | 22<br>(range 4-33 years)<br>n=14 | 13<br>(range 1-64 years)<br>n=31 | NS**<br>(p = 0.06) |
|  | Proportion with PSC | 16.7% | 13.5% | NS*<br>(p=1) |
| **Lesion features** | Lesion spatial distribution | 90% left colorectum<br>10% right colon<br>n=21 | 58% left colorectum<br>42% right colon<br>n=36 | Significant*<br>(p = 0.02) |
|  | Lesion morphology | 33% polypoid<br>22% non-polypoid<br>44% invisible<br>n=18 | 83% polypoid<br>8% non-polypoid<br>8% invisible<br>n=36 | Significant***<br>(p = 0.001) |
|  | Median size of visible lesions (mm) | 45<br>(range 5-50)<br>n=5 | 9<br>(range 2-150)<br>n=25 | Significant***<br>(p = 0.04) |

Where patients had multiple LGD lesions, we extracted DNA separately from each using epithelial enrichment by laser capture microdissection or needle macrodissection. To examine intra-lesion heterogeneity from the largest LGD lesions, we extracted DNA from multiple regions within the same lesion. In total, we extracted DNA from 164 regions of LGD in the discovery cohort and from 106 regions of LGD in the validation cohort. We successfully performed low pass WGS (lpWGS) on all regions, with median coverage of 0.11x (IQR 0.09 - 0.13x). For each region, we derived a whole-genome copy number profile (see Methods).

### Progressor LGD has a higher CNA burden than non-progressor LGD

We used lpWGS data to quantify the burden of somatic copy number alterations (CNAs) in each LGD region. We noted that lesions which appear to be endoscopically and histologically similar can have strikingly different CNA profiles (Figure 2).

**Figure 2:**
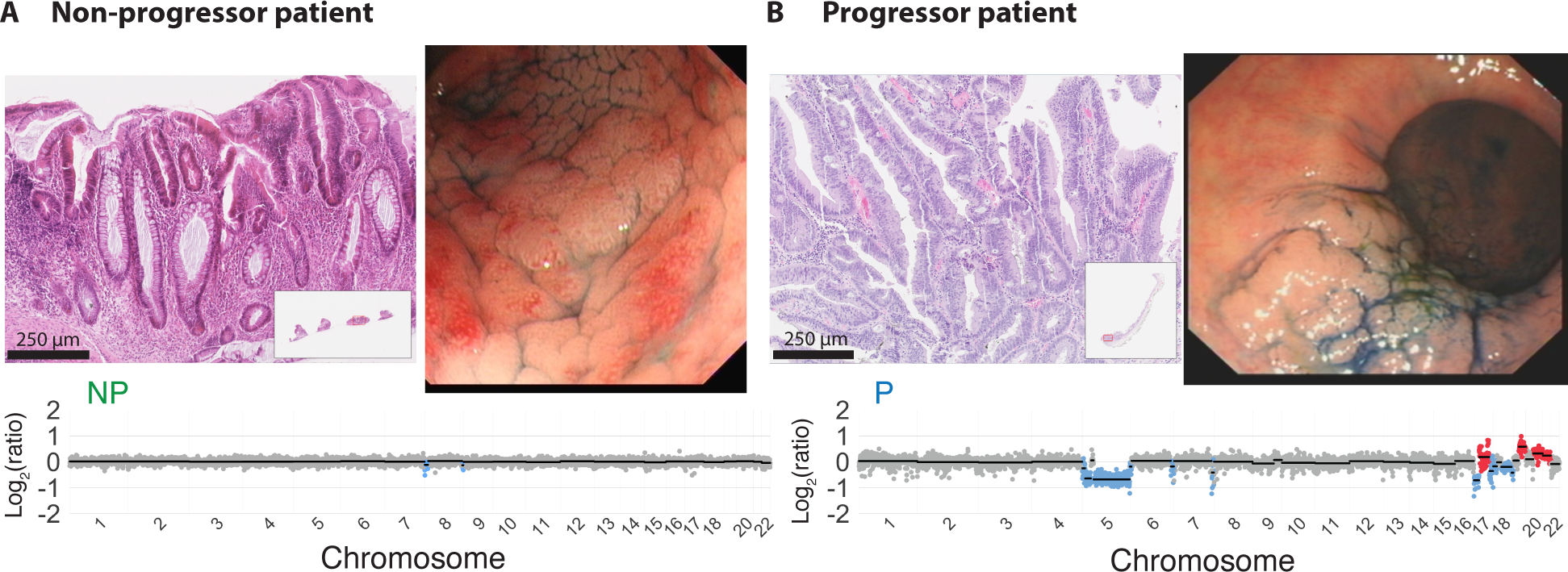
Representative endoscopic and histological images with copy number profiles. Endoscopy images (upper right) and H&E (upper left) with genome-wide copy number profile below for representative non-progressor (**A**) and progressor (**B**) patients.

We compared CNA burden between progressor (P) and non-progressor (NP) lesions and observed a large difference in the number of CNA events between the two groups in the discovery dataset (Figure 3A), and this was highly statistically significant (Figure 3B, median 4 CNAs in NPs vs 16 CNAs in Ps, p=2 x 10^-6^, Mann-Whitney U test). Similarly significant differences were present in the validation cohort (Supplementary Figure S1). We note that for patients who had multiple regions or multiple LGD lesions analysed, we considered only the region with highest CNA burden for this comparison. We compared the burden of CNAs detected after epithelial enrichment by laser capture microdissection versus needle macrodissection and found no significant difference (Supplementary Figure S2).

**Figure 3:**
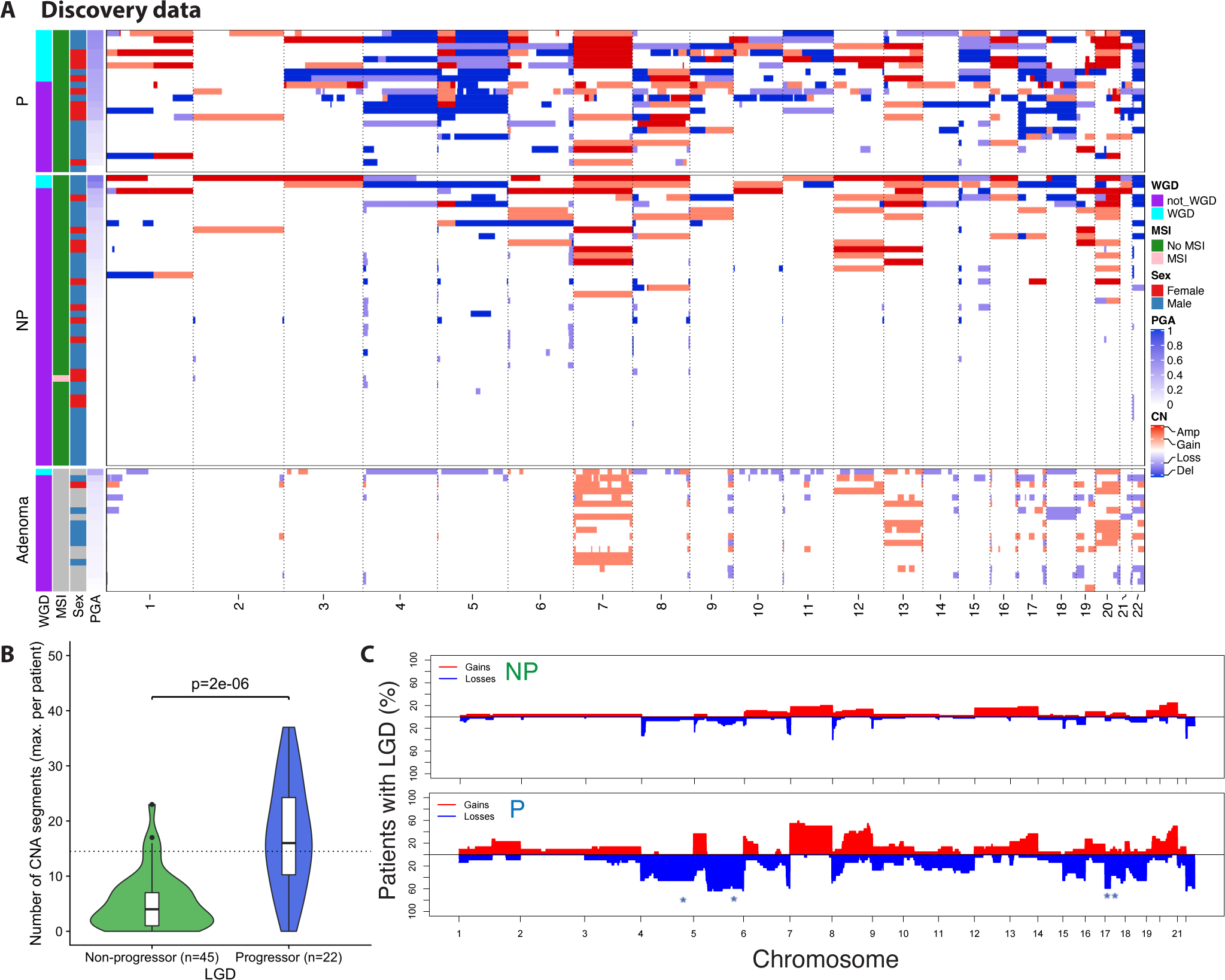
**Genomic alterations in non-progressor versus progressor LGD in discovery cohort. A**. Heatmap of genome-wide copy number alteration for lesions in the discovery LGD cohort, sorted by percent genome altered. Sporadic adenomas included below. **B**. Violin plots showing the number of altered genomic segments in progressor and non-progressor lesions. **C**. Genome-wide CNA frequency for non-progressor (NP, top) and progressor (P, bottom) patients. Stars indicate significant arm level differences at adjusted p-value p<0.01 level. For patients with multi-region analysis the most highly altered sample per patient was included.

We next compared the CNA profiles of our LGD lesions to those of 19 sporadic adenomas from patients without IBD^19^. We found that although there were commonly altered regions (such as gains of chromosome 7, 13 and 20), there were alterations unique or highly enriched in IBD-LGD (Figure 3A).

To determine whether there were specific CNAs associated with progressor LGD, we reviewed the distribution of individual copy number gains and losses across all 67 patients in the discovery dataset (Figure 3C). Evidence of selection is indicated by the non-random distribution of CNA frequencies (assuming a genome wide propensity to generate CNAs) - there were CNAs which were never observed in this cohort (for example, chromosome 4 gains and 2q losses), and there were CNAs that are repeatedly detected (such as loss of chr5q and gain of chr7). Many of the common CNAs found across the progressor cohort were also seen in the non-progressor cohort, albeit at far lower frequency; however, one of the most striking differences between the cohorts was in the frequency of losses involving chromosome 17 (the location of the key tumour suppressor gene *TP53*) which were strongly associated with progressor LGD (Benjamini-Hochberg adjusted p<0.001 for Fisher’s exact test). There were also other specific chromosomal CNAs that were found in a significant proportion of progressor LGDs but not in non-progressor LGDs (Supplementary Table S1). Interestingly, we noted CNA losses involving sub-telomeric segments of chromosomes 4, 5, 6 and 8 in both non-progressors and progressors, either in isolation (as seen in the former) or as part of a chromosomal arm/whole chromosome loss (as seen in latter, Figure 4A).

**Figure 4:**
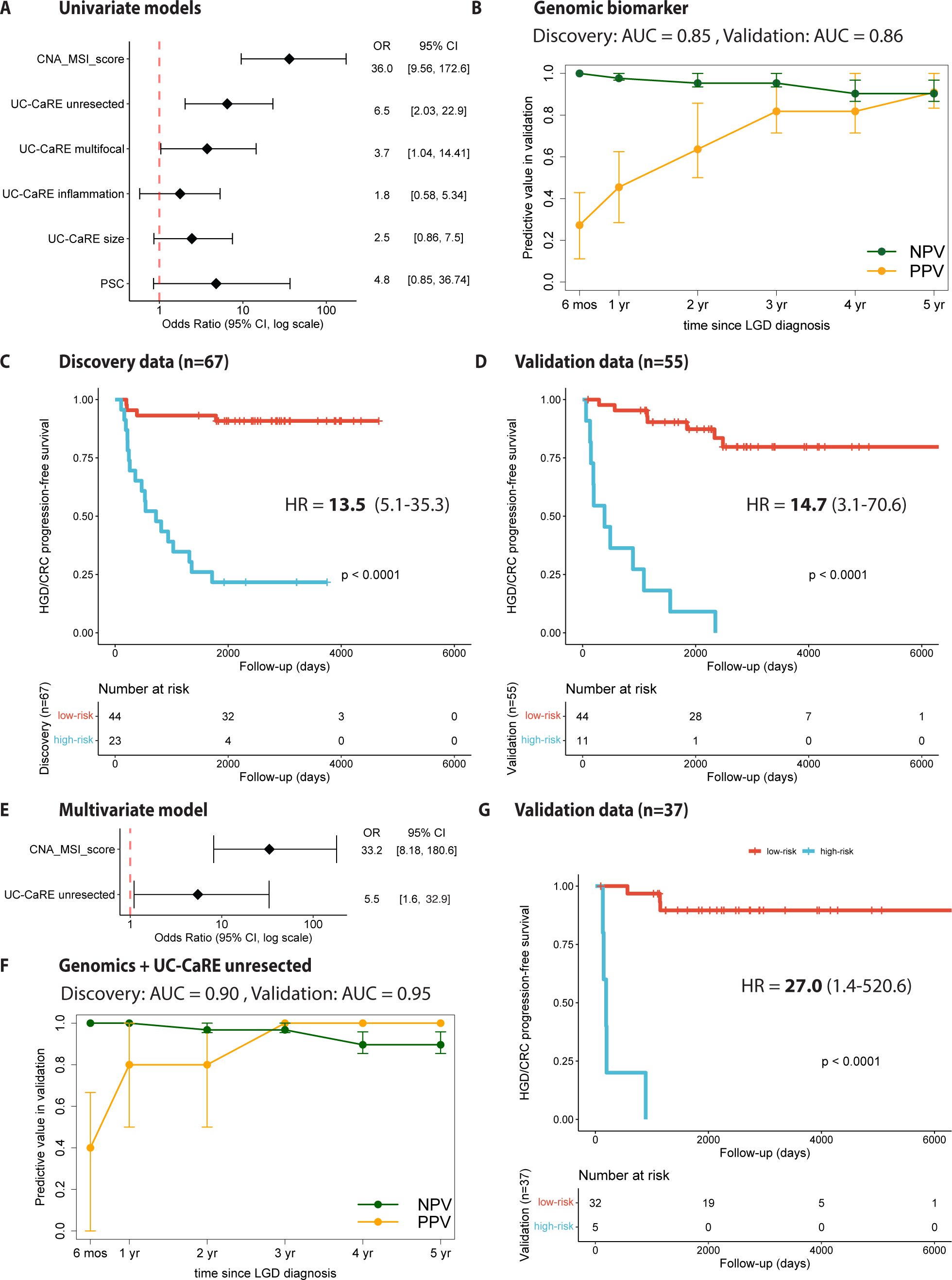
**Model performance for prediction of future HGD/CRC. A**. Odds ratios for univariate models considering genomic score, UC-CaRE, and PSC variables. **B**. AUC values for genomic score alone at 5 years and PPV/NPV over time in validation data with 95% bootstrap confidence intervals. Kaplan-Meier progression-free survival for the discovery (**C**) and validation (**D**) cohorts using a univariate genomic score model. **E**. Odds ratios for multivariate model chosen using stepwise selection. **F**. AUC values for multivariate model at 5 years and PPV/NPV over time in validation data. **G**. Kaplan-Meier curves for validation data; high-risk (>0.5 risk) patients determined by multivariate model prediction.

### Genomic features of LGD can predict future HGD/CRC

We further interrogated the lpWGS data by calling whole genome doubling and microsatellite instability (MSI) status in each region (see Figure 3A). We used lpWGS-derived genomic features (CNA profiles, MSI status) to derive a binary “genomic CNA score” for risk of progression, in which patients are classified as high-risk (score = 1) or low-risk (score = 0). A patient was considered as ‘high-risk’ if 1) the number of CNA segments was > 14.5 (75th percentile of CNA segment number in all discovery data) in any LGD sample sequenced, and/or 2) loss on chromosome 17 in any LGD sample sequenced, and/or 3) any LGD sample was classified as MSI. If none of these applied, then the “genomic CNA score” for that region was zero and the patient would be considered ‘low-risk’.

Univariate logistic regression analysis considering the genomic CNA score and the clinical and endoscopic variables used in UC-CaRE^8^, along with a diagnosis of PSC, found that the genomic CNA score was significantly associated with increased risk of progression to HGD/CRC (odds ratio = 36.0, p= 9e-07; Figure 4A). In the discovery data, the univariate model with genomics alone (CNA score) had an Area Under the receiver operating characteristic Curve (AUC) = 0.85 for accuracy in identifying progressor versus non-progressor patients. When the exact same criteria were applied to data from the independent validation cohort, we found that a univariate CNA score model had an AUC = 0.86 (n=42 patients included in this analysis, see Methods).

Using the genomic CNA score model in the external validation data, we also computed positive predictive value (PPV) and negative predictive value (NPV) over time since the LGD was resected (see Methods). Usually this corresponded to the time of LGD diagnosis, but when this was not available, we used the earliest available time-point with LGD found. We found that PPV = 0.27 at 6 months, PPV = 0.45 at 1 year, and PPV = 0.91 at 5 years post index timepoint (Figure 4B). For patients classified as low-risk, we found that NPV = 1.0 at 6 months, NPV = 0.98 at 1 year, and NPV = 0.90 at 5 years post index timepoint (Figure 4B).

To investigate the utility of lpWGS as a clinical risk stratification tool, we next used Kaplan-Meier survival analysis to compare HGD/CRC progression-free survival curves between low-risk and high-risk patients as determined by the CNA score. In the discovery cohort, patients deemed ‘high-risk’ were significantly more likely to progress to HGD/CRC than patients deemed ‘low-risk’, with a hazard ratio (HR) of 13.5 (log-rank p < 0.0001, Figure 4C). Similarly in the validation cohort, we found that our genomic CNA score effectively stratifies high-risk versus low-risk patients with HR = 14.7 (log-rank p < 0.0001, Figure 4D).

### A multivariate model combining genomic CNA score and clinical information increases predictive power

To determine a parsimonious multivariate model that combined genomics with candidate clinicopathologic variables, we performed stepwise selection using all variables shown in Figure 4A for the discovery data. The best-performing model included our genomic CNA score and the UC-CaRE binary variable of incomplete LGD resection (yes or no). Both variables remained significant in multivariate logistic regression although the odds ratio for our genomic CNA score was 6 times higher than the odds ratio for incomplete resection (Figure 4E). In the discovery data, this multivariate model had an AUC = 0.90 for accuracy in classifying progressor versus non-progressor patients. Here we used predicted risk from the multivariate model and considered a patient to be ‘high-risk’ if their predicted risk was greater than 0.5 and ‘low-risk’ otherwise. When the exact same risk model was applied to the validation data, we found excellent model accuracy with an AUC = 0.95 (n = 29 patients included in this analysis). Using this risk prediction model in the external validation data, we found that PPV = 0.4 at 6 months, PPV = 0.8 at 1 year, and PPV = 1.0 at 5 years post index timepoint (Figure 4F). For patients classified as low-risk, we found that NPV = 1.0 at 6 months, NPV = 1.0 at 1 year, and NPV = 0.90 at 5 years post index timepoint (Figure 4F).

Finally, we used Kaplan-Meier survival analysis to compare HGD/CRC progression-free survival curves between low-risk and high-risk patients as determined by the combined risk model. The risk stratification in the discovery data was identical to the univariate model results shown in Figure 4C. However, when applying to the independent validation data, we found that the combined risk model improves the stratification of high-risk versus low-risk patients in their progression risk with HR = 27.0 (log-rank p < 0.0001, Figure 4G).

### Increased intra-lesion genomic heterogeneity in progressor LGD lesions compared to non-progressor patients

To characterise intra-lesion heterogeneity, we compared the CNA profiles of multi-region sequenced LGD lesions. We derived a CNA-based phylogenetic tree for each multi-region sequenced lesion (median 3 regions per lesion, range 2-8 regions per lesion). As previously described, we saw greater CNA burden in progressor LGD versus non-progressor LGD (Figure 5A-B).

**Figure 5:**
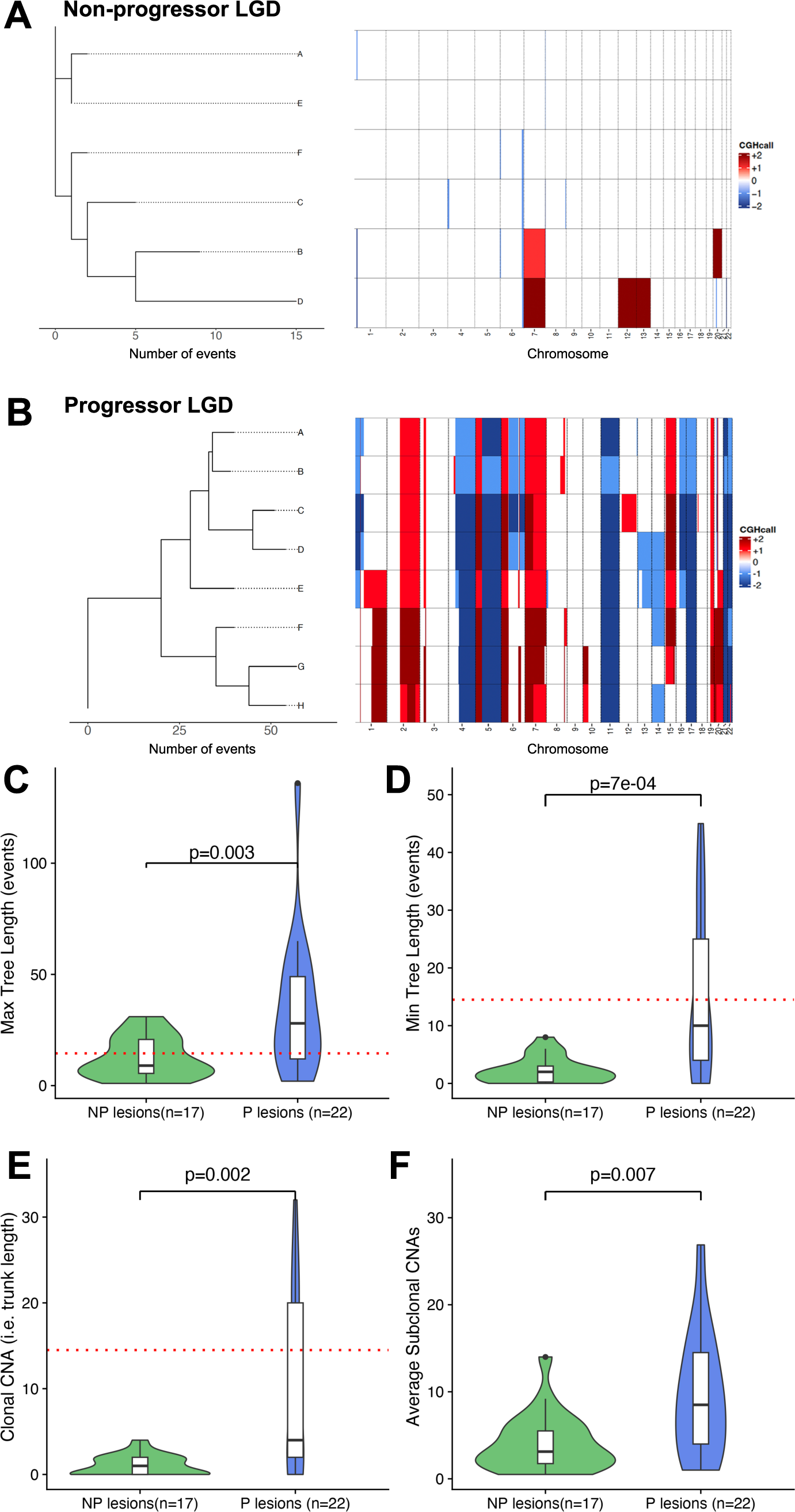
Phylogenetic analysis of multi-region data. Phylogenetic trees (left) and matched copy number profiles (right) for (**A**) a representative non-progressor lesion and (**B**) a representative progressor lesion. Tree statistics for lesions in the discovery cohort that had at least 2 regions sequenced were maximum tree length (**C**), minimum tree length (**D**), clonal CNA (**E**), and average subclonal CNA (**F**).

We measured four tree statistics to compare all progressor (n=17) and non-progressor (n=22) lesion trees (Figure 5C-F). Maximum and minimum tree length (the number of events from a diploid state to the most and least aberrant sample) was significantly higher in progressors compared with non-progressors (Figure 5C,D). The maximum and minimum tree length was 15 to 2 for the non-progressor example (Figure 5A) and 57 to 39 for the progressor example (Figure 5B). Average subclonal CNA (i.e. average branch length from the trunk to the tips) was also significantly higher in progressor legions implying greater ongoing genomic “evolvability” (Figure 5F).

Importantly, the clonal CNA (i.e. trunk length) was much higher in progressor lesions (Figure 5E, and seen when comparing Figure 5A to 5B). Therefore, the detection of an elevated CNA burden in progressor lesions was expected to be robust to intra-lesion heterogeneity, despite the elevated heterogeneity in progressor lesions. To investigate this further, we repeated our analysis of the predictive power of our genomic CNA score, this time choosing a random sample per patient in the validation data rather than the most highly altered region. We calculated similar PPV and NPV values over time since the initial timepoint, confirming that the predictor is robust to lesion heterogeneity (Supplementary Figure S3).

## Discussion

Colorectal cancer (CRC) risk is chronically under- and over-diagnosed in colitis patients, due to lack of a sufficiently accurate predictor of future risk. Here we have found that the pattern and burden of somatic copy number alterations within low-grade dysplasia (LGD) lesions very strongly predicts the future risk of CRC, with positive and negative predictive values at 5 years above 90%. Further, genomic analysis by lpWGS is the single most significant predictor of progression, exceeding the predictive value of any currently used single clinical-pathological feature. We consider this level of predictive power to be clinically useful to support risk-informed decision making in this patient cohort.

Our data is suggestive of a central role for CNA accumulation in the molecular pathogenesis of CA-CRC, that is distinct from the evolutionary trajectory of sporadic colorectal carcinogenesis^11,21–25^. Specific aneuploidies presumably provide a fitness benefit in the highly inflamed colitic microenvironment. For example, while losses involving 17p and gains in 7p are common in sporadic adenomas and IBD-LGD, other events such as 4q loss and 8q gain are significantly over-represented in IBD dysplasia, and chr17 losses are enriched in progressive colitis lesions.

Intra-tumour heterogeneity is an adverse risk factor for progression in a wide variety of cancers^26^. Clonality of premalignant colonic lesions remains an area of interest that is not as well investigated. Advanced sporadic colonic adenomas may be more likely to have sub-clonal CNAs compared to established CRC^27,28^. In a subset of our larger visible lesions where multi-region sequencing was possible, here we observed that “progressor” LGD lesions had greater intra-lesion diversity in keeping with the hypothesis that the elevated cancer risk in these lesions was because of their greater “evolvability”.

In conclusion, we have derived a novel genomic biomarker of future colorectal cancer risk in IBD patients with LGD. This biomarker has major translational potential, and we anticipate it will improve the clinical management of IBD, reduce colorectal cancer diagnoses and improve quality of life for IBD patients.

## Methods

### Human tissue & anonymised data acquisition

FFPE human tissue samples, with corresponding anonymised clinical data, were obtained with ethical approval from St. Mark’s Hospital (NHS REC reference 18/LO/2051) from patients with extensive ulcerative colitis (Montreal Classification E3). All pathology was confirmed by two blinded GI pathologists (Drs MM, MJ, MRJ, and Prof Sir NW) using the corresponding H&E slide. Where two pathologists were in disagreement on dysplasia grading, the review of a third pathologist was sought, and the majority grading severity was chosen.

Where a patient had multifocal LGD, all dysplastic lesions were taken forward for analysis. In our St. Mark’s cohort (called the discovery dataset throughout), we aimed for 1:2 patient matching, with one progressor (P) for every two non-progressors (NP). Clinically relevant risk factors such as patient age, gender, duration of IBD diagnosed, and the presence of PSC (primary sclerosing cholangitis) were matched between P and NP. Moreover, endoscopic factors such as lesion size, shape and location were noted. Censor time is defined as the number of days from the procedure at which the LGD tissue sample is obtained until: 1) the date of the colonoscopy or operation at which either HGD or CRC is first detected (for P), or 2) the date of the final colonoscopy confirming that the patient remains HGD and CRC-free (for NP).

For the Newcastle and Oxford validation cohorts, we replicated this as closely as possible, however these cohorts had only 3 years of follow up data at the censoring time.

### DNA extraction from FFPE tissue

Lesions with low epithelial content were sectioned onto UV-treated, PEN-membrane covered slides (Zeiss) to allow for epithelial enrichment by laser-capture microdissection, or they were sectioned onto charged glass slides for epithelial enrichment by needle macrodissection. In each case six sequential 10 micron sections were used. Sections were dewaxed in xylene for 5 minutes, and rehydrated using 5 minute incubations in dilutions of ethanol (100%, 90%, and 70%) before staining with methyl green (Sigma Aldrich). Sections were briefly washed in water. Sections on PEN-membrane slides were dehydrated in 100% ethanol for 2 minutes. A sequential annotated H&E was used as a guide for epithelial enrichment, either through targeted needle macrodissection or laser capture microdissection with the PALM MicroBeam (Carl Zeiss Microscopy).

DNA was extracted using a modified protocol of the High Pure FFPET DNA Isolation Kit (Roche Life Science). Briefly, the isolated epithelium enriched tissue samples were digested in 23µL proteinase K and 60µL lysis buffer for 16 hours at 56C, followed by enzyme denaturation using a 1 hour incubation at 90C. After cooling to room temperature, 67µL of DNA binding buffer is added and incubated for 10 minutes at room temperature. 33µL of isopropranol was then added and the solution passed through DNA-binding spin columns twice by centrifugation. After three washes of the spin columns using the supplied wash buffers, DNA was eluted in 30µL nuclease-free water with an average yield of about 5ng.

### Library preparation

Between 2-3ng of input DNA was used for library preparation with the NEBNext Ultra II DNA Library Prep Kit for Illumina (New England BioLabs). Minor modifications to the manufacturer’s protocol included the halving of all recommended reagent volumes for cost-effectiveness, the elimination of DNA fragment size selection step prior to addition of adaptor, and optimised dilution of Illumina adaptor (dependent on input DNA). Libraries were barcoded using the NEBNext Multiplex Oligos for Illumina kit (Dual index Primers Set 1), and subjected to 11-13 cycles of library amplification. The final library quantity was confirmed with the Qubit Fluorometer, and quality (fragment size distribution and the presence of unbound adaptor) were verified using the Agilent High Sensitivity D1000 DNA TapeStation system. Libraries were sequenced if the peak fragment size was greater than 184 base pairs. Samples were equimolar pooled and sequenced to a target 0.1x coverage using the Illumina NextSeq 500.

### Low pass whole genome sequencing – pre-processing bioinformatics pipeline

Bioinformatic analysis was performed on the QMUL High Performance Computing Cluster (Apocrita^29^). After merging flow cell FASTQ output for every sample, reads in the generated files are subjected to adaptor trimming using Skewer^30^, with trimmed reads less than 35 base pairs long filtered out. A quality control report (including per base sequence quality and estimated duplicate rate) is generated for each FASTQ file using FASTQC^31^. The reads are aligned against the reference genome hg19 using the Burrows-Wheeler Aligner package BWA-MEM^32^ to produce corresponding SAM files. These generated files are then converted to compressed BAM files, with filtering out of poorly mapped reads (defined as a MAPQ score of less than 10^-37/10^, which is approximately less than 1 in 5000) using SAMtools^33^. Duplicate reads were subsequently filtered out using Picard Tools^34^, and BAM index files created for more rapid data extraction including read visualisation on Integrated Genome Viewer^35^. A quality control report relating to mapping quality is generated for each BAM file using BAMQC^36^, and mean genome coverage estimated using GATK^37^.

Next, the QDNAseq^38^ package in R was used to arrange BAM reads into bins that are 500 kilobase pairs (kbp) in size, followed by correction for GC content, the filtering out of “blacklisted” bin clusters identified by ENCODE^39^ (poorly mapped genomic regions rich in nucleotide repeats such as centromeres, telomeres and microsatellites) and by the QDNAseq package developers and our own team (other recurrently deleted bin clusters seen in both normal and neoplastic tissue, often located in peri-centromeric and sub-telomeric regions), and then the smoothing of outliers. Sex chromosomes are also excluded from downstream analysis. The remaining 4,401 bins are subsequently normalised against the mean followed by log2 transformation and segmented using R package DNAcopy^40^. To allow for differences in cellularity and ploidy, we then used R package ACE^41^ to obtain a best fit model of the segmented copy numbers to determine the cellularity and ploidy for each sample with the following function call: squaremodel(template, penalty = 0.5, penploidy = 0.5), where template is the segmented copy numbers output from QDNAseq. Chromosomal copy number alterations were then identified using CGHcall^42^ where per sample cellularity was given as input from ACE output, with each bin assigned a probability for the different types of copy number alteration (losses, gains, complete deletions and amplifications of chromosomal regions). Bins with a copy number alteration probability of greater than or equal to 50% probability are “called” as per CGHcall recommendations and taken forward for downstream analysis in R (see Figure 3 for representative copy number profiles). Copy number alterations were also derived from sporadic (non-IBD) adenomas as previously described^19^.

### Low pass whole genome sequencing – downstream analysis of copy number

The following measurements were made to compare the tissue types of interest:

1. Genomic aneuploidy: the proportion of the 500kbp bins with a called copy number alteration.
2. The number of chromosomal CNA events: the number of individual segments where the constituent bins have a called copy number alteration (e.g. in Figure 2, part b there are 2 CNA events on chromosomes 17).
3. A chromosomal arm loss or gain of interest: a chromosomal arm gain or loss of interest was considered to be present if a segment(s) within that arm have been annotated to have a gain, loss, deletion or amplification by CGHcall.

Statistical comparisons of these measurements were made on R using Mann-Whitney U tests (for measurements 1-2) or Fisher’s exact tests (measurement 3), with a p-value of 0.05 used as a cut- off for significance. Where multiple comparisons are made, a Benjamini-Hochberg correction was used to generate adjusted p-values. Violin plot generation was performed in R using the package ggplot2.

### MSI classification pipeline using low pass whole genome sequencing

In this study, we employed our in-house developed method MILO (Microsatellite Instability in Low-quality biopsies) to investigate the MSI status in all IBD biopsies. MILO is specifically designed for the assessment of MSI status in low-quality samples, including low-coverage FFPE biospecimens. We identified highly sensitive genomic features for MSI classification from publicly available MSI cohorts with low-quality data and subsequently utilised this information to create MSI classifiers, referred to as MILO. We demonstrated the performance of MILO using previously labelled datasets, achieving an accuracy of 0.99. For more comprehensive information, please refer to the original manuscript.

### Whole genome doubling classification

Using International Cancer Genome Consortium (ICGC) segment calls^43^ we binned the genome into 4Mb bins to mimic data expected in low coverage WGS. Here we took the median calls for each bin when comparing overlapping segments. For each sample we calculated the number of segments after binning by detecting changes in call status in adjacent bins across the genome. We calculated percent genome altered (PGA) by measuring the fraction of bins across the genome that did not equal the median copy number of the bins of the sample (baseline copy number). Taking the called genome-doubled (GD) status per sample from the ICGC resource, we trained a support vector machine using the e1071 package in R (no scaling, a linear kernel and a cost of constraints violation of 5). The classifier applied to these data calls GD as present in a sample (Figure 3) if:

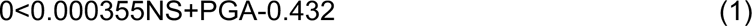

(Where NS denotes the number of segments, PGA the percentage genome altered [fraction of genome with non-baseline copy number]). On Pan-Cancer Analysis of Whole Genomes (PCAWG) data, this classifier calls 83.8% of GD samples correctly, and 93.2% of non-GD correctly^19^.

### Prediction accuracy, multi-region analysis and phylogenetic tree reconstruction

For patients with samples taken from multiple regions and/or at multiple time points, the region with maximal number of chromosomal CNA segments at the first time point was used in the calculation of the number of CNA segments and taken forward for survival analysis. Patients with LGD were censored when HGD or CRC was first detected, or at the final recorded clinical report if the patient did not progress to HGD or CRC. For consistency in the calculations of Area Under the Curve (AUC) using the ROCR^44^ package in R, we utilised all patients in the discovery data and those patients in the validation data who either progressed to HGD or CRC in the first 5 years of follow-up from the initial timepoint (‘progressors’) or remained HGD/CRC-free for at least 5 years of follow-up (‘non-progressors’). For time-dependent NPV and PPV curves, 95% bootstrap confidence intervals were calculated by simulating 1000 runs with ¼ of validation data held-out and then calculating NPV and PPV for each timepoint.

To assess intra-lesion heterogeneity, MEDICC2^45^ was used to create phylogenetic trees for lesions with multi-region samples, where branch lengths are proportional to copy number ’events’. The number of “clonal” CNAs can be visually interpreted as the ‘trunk length’ of the phylogenetic tree, while sub-clonal CNAs per sample are the average distance of each tip from the trunk.

### Logistic regression modelling and survival analyses

For the case-control study design, univariate and multivariate logistic regression analyses were performed for the discovery data in R using the *glm* function. The final multivariate model was chosen by performing backward stepwise selection in R using the *step* function and initially including all UC-CaRE variables and genomic score. For classification of “low-risk” and “high-risk” groups using the multivariate model, the predicted probabilities from function *glm.predict* in R were utilised and a threshold of 0.50 was used to designate a high-risk patient. Kaplan-Meier (KM) survival estimation analyses with right censoring were performed in R using the survival package and plotting of KM curves was performed using the survminer package. Log-rank tests were used for statistical comparison of the two KM curves.

### Data availability

Processed data needed to recreate all results will be available in the Supplementary. Copy number segmentations and calls for all 270 samples will be made available on Mendeley. Raw sequencing data is not available due to the retrospective and anonymous nature of the study.

### Code availability

Computer code for reproducing results will be made available on Github.

## Acknowledgements

The study was principally funded by Cancer Research UK (Early Detection project award 25901) to TA Graham and SJ Leedham, and the Barts Charity (large project grant 472-2300) to TA Graham. We acknowledge a contribution from Bowel Research UK to TA Graham via their PhD student funding scheme, and pilot funding from the 40tude charity to TA Graham and AL Hart. TA Graham received additional funding from Cancer Research UK (CRUK A19771 and DRCNPG- May21_100001). SJ Leedham was supported by CRUK Program Grant (DRCNPG- Jun22\100002). I Al Bakir was funded by an MRC clinical studentship. Support for this research was provided by the NIHR Imperial BRC. NA Wright received funding from Cancer Research UK Program Grant A2187. M Jansen is supported by a Cancer Research UK Clinician Scientist Fellowship (A22745). K Curtius received funding from an MRC HDR-UK programme (UKRI Rutherford Fund Fellowship). This work was supported by AGA Research Foundation (AGA Research Scholar Award AGA2022-13-05; K Curtius) and NIH grants (R01 CA270235, P30 CA023100; K Curtius). This work was supported in part by a Merit Review Award I01 BX005958 (K Curtius) from the United States (U.S.) Department of Veterans Affairs Biomedical Laboratory Research and Development Service. The contents do not represent the views of the U.S. Department of Veterans Affairs or the United States Government. The study was supported in part by the NIDDK-funded San Diego Digestive Diseases Research Center (P30 DK120515). JE East is funded by the National Institute for Health Research Oxford Biomedical Research Centre. The views expressed are those of the author and not necessarily those of the National Health Service, the National Institute for Health Research, or the Department of Health. The authors would like to thank Dr. Adam Brentnall and Dr. Ronghui Xu for helpful statistical discussions.

