## Supplementary Information for "Low coverage whole genome sequencing of low-grade dysplasia strongly predicts colorectal cancer risk in ulcerative colitis"

**Supplementary Figure S1: Genomic alterations in non-progressor versus progressor LGD in validation cohort. A**. Heatmap of genome-wide copy number alteration for lesions in the validation LGD cohort, sorted by percent genome altered. **B**. Violin plots showing the number of altered genomic segments in progressor and non-progressor lesions. **C**. Genome-wide CNA frequency for non-progressor (NP, top) and progressor (P, bottom) patients. For patients with multi-region analysis the most highly altered sample per patient was included.

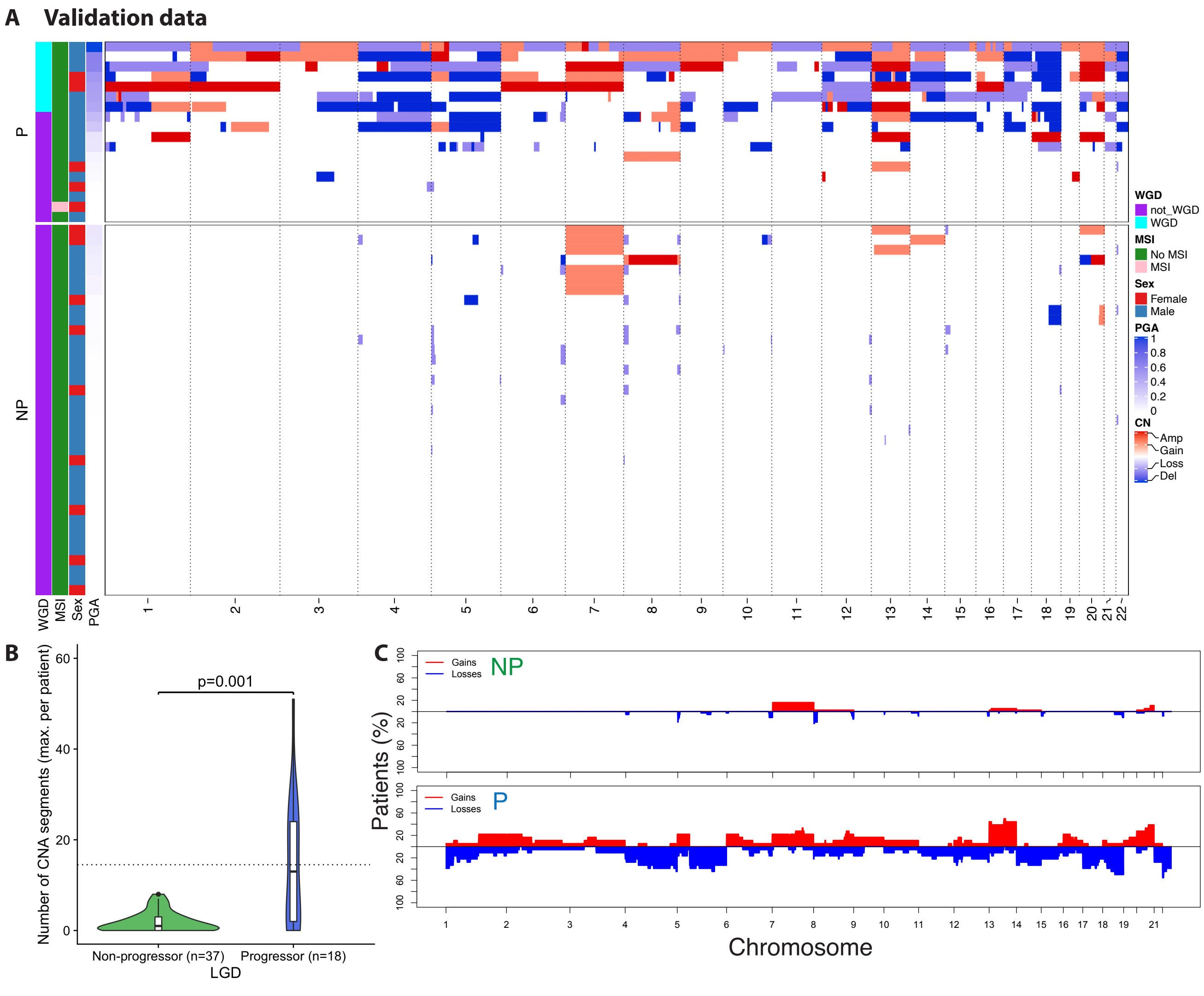

**Supplementary Figure S2: Detection of copy number altered (CNA) segments by epithelial isolation method.** Laser capture microdissection (LCM) tissue (blue) vs. Scrape tissue (pink). For LCM n = 76, for Scrape n = 46. By Mann-Whitney U tests for strategies of P vs P and NP vs NP groups, the only statistically significant difference here was LCM NP (median 3.5 CNA segments) vs Scrape NP groups (median 1 CNA segment) with p=2.9e-5.

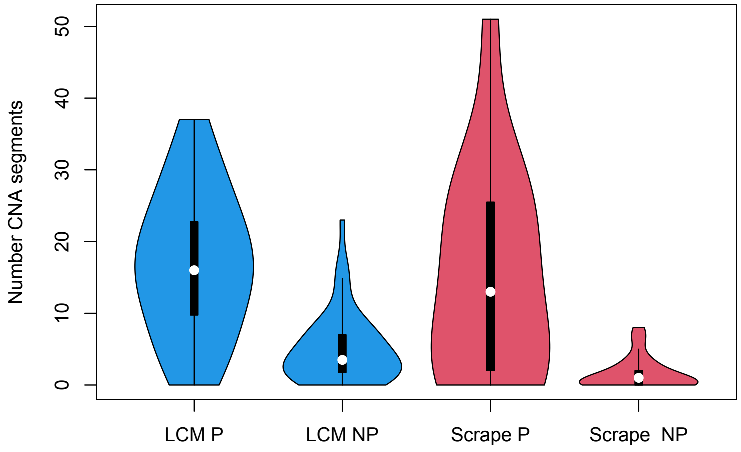


**Supplementary Figure S3: Predictive accuracy of genomic biomarker using a randomly selected sample from each patient.** Re-calculation of PPV and NPV of genomic CNA score biomarker when a random sample for each patient in the validation cohort is used for prediction. 95% confidence intervals are shown for each predicted time point (1000 simulations of random sample selection). PPV= positive predictive value. NPV = negative predictive value.

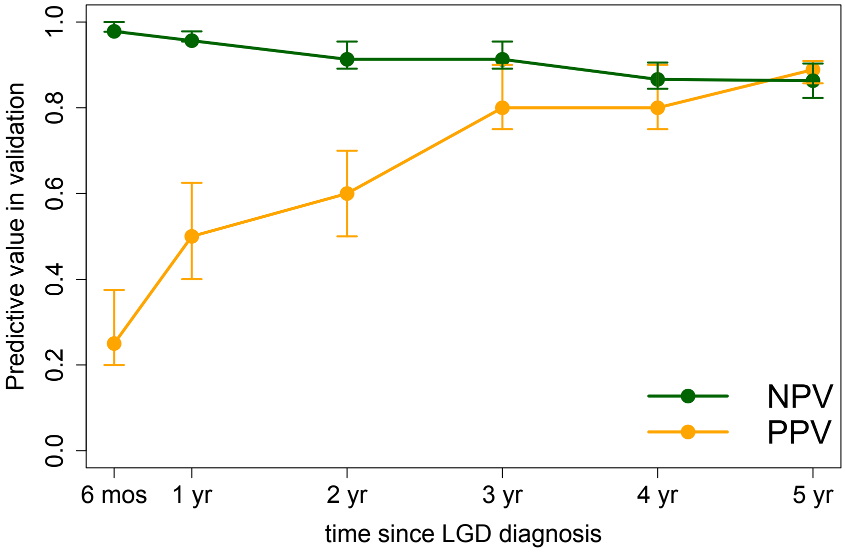

**Supplementary Table S1: Top chromosome arm level differences in max frequency of changes across samples in progressor vs non-progressor patients**. *Adjusted p-values provided for Fisher's exact test with Benjamini-Hochberg multiple hypothesis testing correction applied.

| Chromosomal arm with CNA | Max frequency in progressors (n = 22) | Max frequency of non-progressors (n = 45) | Odds ratio | Adjusted Fisher's exact test p-value* |
|---|---|---|---|---|
| **Loss on 4q** | 0.45 | 0.07 | 11.14 | 0.008 |
| **Gain on 5p** | 0.36 | 0.04 | 11.74 | 0.012 |
| **Loss on 5q** | 0.64 | 0.16 | 9.09 | 0.004 |
| **Gain on 7p** | 0.59 | 0.18 | 6.45 | 0.012 |
| **Gain on 8q** | 0.55 | 0.13 | 7.50 | 0.011 |
| **Loss on 11p** | 0.27 | 0 | Inf | 0.011 |
| **Loss on 11q** | 0.32 | 0.02 | 19.51 | 0.012 |
| **Loss on 17p** | 0.59 | 0.07 | 18.96 | <0.001 |
| **Loss on 17q** | 0.5 | 0.02 | 40.96 | <0.001 |
| **Loss on 18q** | 0.45 | 0.09 | 8.21 | 0.012 |
